# Airway microbiota in young people across four continents differ by country, asthma status and inflammatory phenotype

**DOI:** 10.1101/2025.03.18.25324207

**Authors:** Steven L. Taylor, Collin R. Brooks, Lucy Pembrey, Sarah K. Manning, Levi Elms, Harriet Mpairwe, Camila A Figueiredo, Aida Y Oviedo, Martha Chico, Jeroen Burmanje, Hajar Ali, Irene Nambuya, Pius Tumwesige, Steven Robertson, Charlotte E. Rutter, Karin van Veldhoven, Susan Ring, Mauricio L. Barreto, Philip J. Cooper, Alvaro A. Cruz, Neil Pearce, Geraint B. Rogers, Jeroen Douwes, the WASP Study Group

**Affiliations:** Microbiome and Host Health Programme, South Australian Health and Medical Research Institute, Adelaide, Australia; College of Medicine and Public Health, Flinders University, Bedford Park, Australia; Research Centre for Hauora and Health, Massey University, Wellington, New Zealand; London School of Hygiene & Tropical Medicine (LSHTM), London, UK; MRC/UVRI and LSHTM Uganda Research Unit, Entebbe, Uganda; Federal University of Bahia, Salvador, Brazil; Fundación Ecuatoriana Para Investigación en Salud, Quito, Ecuador; Department of Non-communicable Disease Epidemiology, LSHTM, London, UK; Population Health Sciences, Bristol Medical School, University of Bristol, Bristol, UK; MRC Integrative Epidemiology Unit at University of Bristol, Bristol, UK; Center for Data and Knowledge Integration for Health, Fiocruz, Bahia, Brazil; School of Medicine, Universidad Internacional del Ecuador, Quito, Ecuador; Institute of Infection and Immunity, St George’s University of London, London, UK; ProAR, Federal University of Bahia, Salvador, Brazil

**Author notes:** **Corresponding author:** Dr. Steven Taylor, Address: North Terrace, South Australian Health and Medical Research Institute, Adelaide, Australia, 5000. Contributed equally (Geraint B. Rogers, Jeroen Douwes).

**Keywords:** Kew Words: Brazil, Ecuador, Uganda, Africa, South America, childhood asthma, *Haemophilus influenzae*, neutrophil, ALSPAC

## Abstract

**Background:** Asthma is an umbrella diagnosis encompassing distinct pathophysiological mechanisms. While a global problem, our understanding of the interplay between respiratory microbiology and airway inflammation is largely from populations in high income settings. As a result, treatment approaches align poorly with asthma characteristics in less studied populations.

**Objective:** To identify conserved and geographically distinct relationships between airway inflammation and microbiota characteristics in young people with asthma.

**Methods:** We conducted a cross-sectional study performing inflammatory phenotyping, microbiota analysis, and enumeration of total bacteria, *Haemophilus influenzae* and *Moraxella catarrhalis* on 488 induced sputum samples from Brazil (n=68), Ecuador (n=119), Uganda (n=69), New Zealand (n=187), and the United Kingdom (n=45). Microbiological characteristics were compared by country, asthma status, and inflammatory characteristics, adjusting for age and sex.

**Results:** Inflammatory phenotypes and airway microbiota differed between countries, with Uganda characterised by greater neutrophil%, microbial diversity, and bacterial load. Across all countries, microbiota similarity differed by asthma status (*P*=0.012). Within participants with asthma, microbiota similarity for neutrophilic and eosinophilic phenotypes differed from paucigranulocytic (*P*<0.001 and *P*=0.020, respectively) and from each other (*P*<0.001). Neutrophil% was strongly associated with microbiota composition (*P*<0.001) and positively associated with bacterial load and opportunistic pathogens (*P*<0.05). In contrast, eosinophil% was less strongly associated with microbiota similarity (*P*=0.033), positively associated with *Streptococcus* (*P*=0.0009), but not associated with bacterial load (*P=*0.787). Country-specific associations between sputum inflammation and microbiology were evident.

**Conclusion:** Both airway inflammation and microbiology varied geographically in young people with asthma. Associations between microbiota characteristics and neutrophilic phenotype were conserved.

**Key messages:** *What is already known on this topic:*

- Asthma treatment response and severity are associated with airway inflammation and microbiology.
- Most asthma research is performed in high income countries and the generalisability in other settings is unclear.

*What this study adds:*

- Asthma inflammatory phenotypes and airway microbiota vary across high income (New Zealand and the United Kingdom) and low to middle income (Brazil, Ecuador, Uganda) countries.
- The association between airway microbiota and neutrophilic and eosinophilic inflammation is complex and varied between countries.

*How this study might affect research, practice or policy:*

- Understanding variation in underlying pathophysiology between countries can inform improved deployment of maintenance asthma therapies, such as macrolides and inhaled corticosteroids, that target specific inflammatory pathways.

## INTRODUCTION

Asthma is associated with several distinct pathophysiological presentations (deemed inflammatory phenotypes), including type 2 (T2)-high/eosinophilic and neutrophilic disease. Different inflammatory phenotypes are associated with variations in treatment response and provide a basis for targeted treatment strategies. For example, inhaled corticosteroid (ICS) therapy is typically effective for those with early-onset, eosinophilic asthma (EA), but provides limited benefit in those with late-onset EA, non-T2-high (T2-Low) or neutrophilic asthma (NA) [1].

Airway microbiology varies with asthma inflammatory phenotypes [2–5], likely reflecting differences in airway pathophysiology and underlying aetiology. For example, lower microbiota diversity and increased prevalence of opportunistic pathogens, including *Haemophilus influenzae* and *Moraxella catarrhalis*, have been associated with NA [2–6]. Airway microbiology has been associated with differential response to macrolide maintenance therapy in adults with poorly-controlled asthma [7], outperforming endotype classification or inflammatory cell-based prediction of treatment response [8].

While the distribution of asthma subtypes is now well-documented within some populations, particularly those in high income countries (HICs) [9–12], far less is known about asthma characteristics in low and middle-income countries (LMICs). Recently, we reported considerable variance in asthma phenotypes between Brazil, Ecuador, New Zealand, Uganda, and the United Kingdom [9], suggesting that factors, such as environmental exposures or socioeconomic status, may influence patterns of asthma pathophysiology. Although no other study has examined asthma phenotype prevalence comparing HICs and LMICs on the basis of airway inflammation, our observations are consistent with the earlier findings of the International Study of Asthma and Allergic diseases in Childhood (ISAAC). Specifically, ISAAC reported that the proportion of asthma attributable to atopy differs between countries, with an average population attributable fraction of 46% observed in HICs and 20% in LMICs [13].

The extent to which geographic variation in asthma phenotype prevalence is reflected in differences in airway microbiology, and how any such differences relate to treatment outcomes and asthma-associated complications, remains unclear. In this component of the World ASthma Phenotypes (WASP) study, termed WASP-biome, we assessed the airway microbiota characteristics of children and young adults with and without asthma in Brazil, Ecuador, New Zealand, Uganda and the UK. Our principal aim was to identify conserved and geographically distinct relationships between inflammatory phenotypes and airway microbiota characteristics in young people with asthma.

## METHODS

Detailed methods are provided in the online supplement.

### Study design

The WASP study is a cross-sectional observational investigation of asthma phenotypes in young people in HICs and LMICs [14]. The study was conducted in five centres: Bristol, UK (Avon Longitudinal Study of Parents and Children, ALSPAC) [15–17]; Wellington, New Zealand; Salvador, Brazil; Quininde, Ecuador; and Entebbe, Uganda (**Table 1**) [9]. WASP-biome involves a subgroup of participants for whom a secondary sputum aliquot was available for microbiota analysis. No other selection criteria were defined for inclusion in WASP-biome.

**Table 1:** Study characteristics of young people with asthma.

|  | **Brazil (Salvador)** | **Ecuador (Quininde)** | **Uganda (Entebbe)** | **New Zealand (Wellington)** | **United Kingdom (Bristol)** |
| --- | --- | --- | --- | --- | --- |
| N | 60 | 89 | 61 | 129 | 25 |
| Female, n (%) | 40 (67%) | 41 (46%) | 45 (73.8%) | 55 (42.6%) | 17 (68%) |
| Age (years), median (IQR) | 17.98 (16.4-19.6) | 11.58 (10.79-13.65) | 15.09 (14.01-16.51) | 11.49 (9.97-14.99) | 26.00 (25.58-26.25) |
| BMI (kg/m<sup>2</sup>), median (IQR) | 22.63 (20.5-25.4) | 18.76 (17.33-21.94) | 20.18 (18.26-22.98) | 19.39 (17.08-22.48) | 26.53 (22.68-31.25) |
| ACQ7 score, median (IQR) | 0.67 (0.17-1.50) | 0.00 (0.00-0.00) | 0.83 (0.00-1.67) | 0.86 (0.33-1.29) | 0.67 (0.17-1.17) |
| ACQ7 level, n (%) |  |  |  |  |  |
| Well controlled | 43 (72.9%) | 84 (94.4%) | 41 (67.2%) | 100 (82.0%) | 22 (88.0%) |
| Uncontrolled | 16 (27.1%) | 5 (5.6%) | 20 (32.8%) | 22 (18.0%) | 3 (12.0%) |
| ICS use past 12 months, n (%) | 10 (17.2%) | 6 (10.7%) | 3 (5.17%) | 84 (65.6%) | 15 (75%) |
| SABA use past 12 months, n (%) | 31 (53.5%) | 13 (23.2%) | 19 (31.7%) | 118 (92.2%) | 21 (100%) |
| FEV<sub>1</sub> (% predicted), mean (STD) | 91.47 (10.10) | 98.04 (11.52) | 97.67 (11.05) | 92.93 (13.10) | 97.87 (13.80) |
| FVC (% predicted), mean (STD) | 96.72 (15.08) | 96.36 (10.77) | 101.91 (12.67) | 99.97 (10.94) | 103.69 (13.75) |
| Atopy, n (%) | 51 (85%) | 29 (32.6%) | 31 (51.7%) | 99 (76.7%) | 15 (79%) |
| Inflammatory phenotype, n (%) |  |  |  |  |  |
| Eosinophilic | 19 (31.7%) | 28 (31.5%) | 12 (19.7%) | 64 (49.6%) | 8 (32.0%) |
| Neutrophilic | 3 (5.0%) | 4 (4.5%) | 24 (39.3%) | 9 (7.0%) | 3 (12.0%) |
| Paucigranulocytic | 37 (61.7%) | 52 (58.4%) | 19 (31.1%) | 51 (39.5%) | 13 (52.0%) |
| Mixed granulocytic | 1 (1.7%) | 5 (5.6%) | 6 (9.8%) | 5 (3.9%) | 1 (4.0%) |
| Sputum neutrophils (%), median (IQR) | 5.75 (1.38-15.62) | 10.32 (5.00-22.25) | 60.59 (30.35-85.00) | 20.00 (8.48-44.15) | 28.50 (15.29-46.88) |
| Sputum eosinophils (%), median (IQR) | 1.00 (0.00-4.75) | 0.73 (0.12-4.25) | 0.75 (0.0-3.50) | 3.25 (0.75-9.25) | 1.39 (0.25-3.25) |

IQR: Interquartile range, STD: Standard deviation, BMI: Body mass index, ACQ7: Asthma control questionnaire, ICS: Inhaled corticosteroids, SABA: Short acting beta-agonist, FEV<sub>1</sub>: Forced expiratory volume in 1 second, FVC: Forced vital capacity

This study was approved by the LSHTM ethics committee (ref: 9776). Informed consent was obtained from all participants or their parents/carers before taking part.

### Asthma diagnosis, sputum collection, and inflammatory phenotyping

Asthma was defined across all centres as wheeze or whistling in the chest and/or use of asthma medication in the past 12 months, using the validated ISAAC questionnaire [18–20]. Induced sputum samples were collected using a standardised protocol [21]. Stored, unprocessed induced sputum aliquots were used for microbiota analysis. Inflammatory phenotypes were determined from differential cell counts, performed on freshly collected samples. Samples with ≥30% squamous cells were excluded due to suspected high upper airway contamination. Asthma inflammatory phenotypes were defined as: eosinophilic asthma (EA): ≥2.5% eosinophils and <61% neutrophils; mixed granulocytic asthma (MGA): ≥2.5% eosinophils and ≥61% neutrophils; neutrophilic asthma (NA): <2.5% eosinophils and ≥61% neutrophils; and paucigranulocytic asthma (PGA): <2.5% eosinophils and <61% neutrophils [9].

### Microbiota characterisation

Induced sputum underwent DNA extraction, quantitative PCR (qPCR) enumeration of total bacteria, *Haemophilus influenzae* and *Moraxella catarrhalis*, and 16S rRNA sequencing. Mock sample and blank extraction controls were analysed to assess taxon representation and reagent contaminants, respectively. Reads have been deposited in the European Bioinformatics Institute European Nucleotide Archive (PRJEB77703).

### Data analysis

Analyses were conducted to assess microbiota differences (i.e. β-diversity, α-diversity, bacterial load, and relative abundance of specific taxa) between: 1) different countries (for participants with asthma and without, separately); 2) asthmatics and non-asthmatics (study-wide and within country); 3) asthma inflammatory phenotypes (study-wide and within country) and 4) asthma inflammatory cell percentage (study-wide and within country). All analyses were adjusted for age and sex, with analyses involving multiple countries also adjusted for location. Analysis of airway inflammation was performed based on comparisons between the four inflammatory phenotypes, and with percentage neutrophils (neutrophil%) and eosinophils (eosinophil%), mutually adjusted for each other.

## RESULTS

### Participant characteristics

Of 920 WASP participants, sputum was collected from 658 with asthma and 262 with no asthma. Of these, 488 (asthma n=364, no asthma n=124) provided sufficient, quality sputum for microbiota analysis (WASP-biome). Study participation by centre is shown in **Figure 1A** and WASP-biome participant characteristics are detailed in **Table 1**. WASP-biome sub-group characteristics were consistent with the wider WASP study cohort (**Supplementary Table 2**), with the highest proportion of NA observed in Uganda and the highest proportion of EA in NZ.

**Figure 1:**
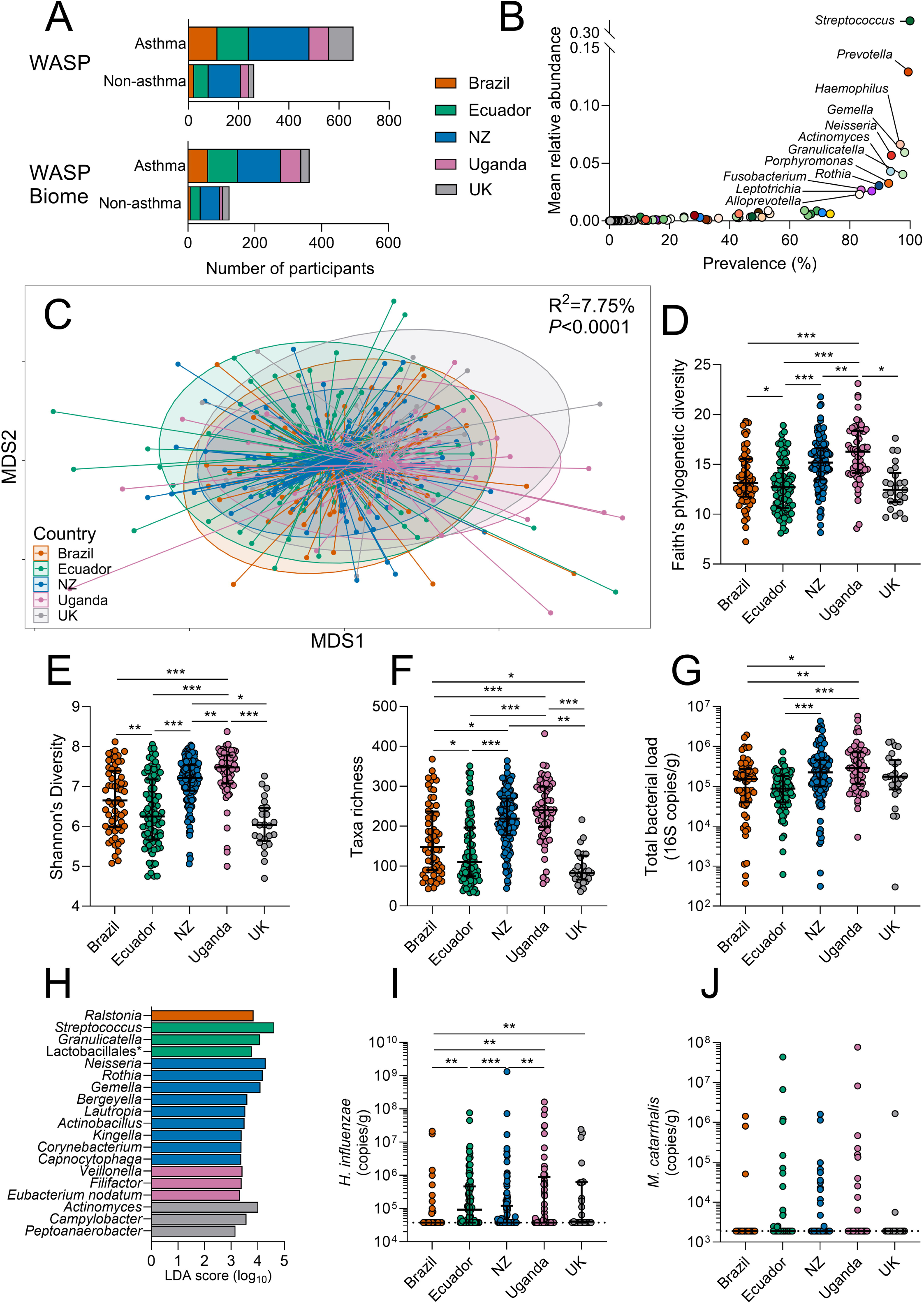
Sputum microbiota differs by country in participants with asthma. A) Overview of sample size for the WASP study and WASP-biome study. B) Taxa summary plot showing the mean relative abundance and prevalence of genera, coloured by phyla (Actinobacteriota: blues, Bacteroidota: oranges, Bacillota: greens, Pseudomonadota: reds, Fusobacteriota: purples, Campilobacterota: yellow, Spirochaetes: brown). C) Nonmetric multidimensional scaling (NMDS) plot of Bray-Curtis dissimilarity. D) Faith’s phylogenetic diversity. E) Shannon’s diversity. F) Taxa richness. G) Total bacterial load derived from qPCR. H) Taxa differentially present by country. I) *Haemophilus influenzae* abundance derived from qPCR. J) *Moraxella catarrhalis* abundance derived from qPCR. Statistics: C) Permutational multivariate analysis of variance including variables: country, age and sex; D-G, I, J) Ordinal logistic regression including variables: country, age and sex; H) Linear discriminant analysis (LDA) effect size, with cut-offs of LDA ≥3 and p<0.05. *** adjusted *P*<0.001, ** adjusted *P*<0.01, * adjusted *P*<0.05.

### Sputum microbiota characteristics differ between countries

A total of 135 bacterial taxa were identified in participants with asthma. Taxa prevalence and mean relative abundance are shown in **Figure 1B**, and variance between centres shown in **Supplementary Figure 1**. Forty-six bacterial taxa were detected in ≥10% of the samples within any of the five centres (**Supplementary Figure 1**). *Streptococcus* was the most commonly detected genus and was present in all samples (mean relative abundance=0.32, STD=0.11), followed by *Prevotella* (prevalence=99.4%), *Gemella* (98.2%)*, Granulicatella* (97.6%)*, Haemophilus* (96.8%)*, Neisseria* (93.8%), *Actinomyces* (93.5%), and *Porphyromonas* (92.9%).

Between-country analysis revealed significant differences in microbiota composition, as defined by Bray-Curtis dissimilarity (R^2^=7.75%, *P*<0.0001, **Figure 1C** and **Supplementary Table 3**). Pairwise comparisons showed the greatest difference between NZ and Uganda (pseudo-F=11.5, **Supplementary Table 3**) and the smallest between Brazil and Ecuador (pseudo-F=2.81). Countries also differed in α-diversity indices (Faith’s phylogenetic diversity, Shannon diversity, and taxonomic richness count) (**Figure 1D-F)** and total bacterial load (**Figure 1G**), with Uganda having both higher α-diversity and bacterial load, while Ecuador and the UK had the lowest. Nineteen bacterial genera displayed significant differences in abundance between countries (**Figure 1H**), with *Ralstonia*, *Streptocococcus*, *Neisseria*, *Veillonella*, and *Actinomyces* the highest in Brazil, Ecuador, NZ, Uganda, and the UK, respectively. Assessment of the absolute abundance of the Gram-negative respiratory pathogens, *H. influenzae* (**Figure 1I**) and *M. catarrhalis* (**Figure 1J**), revealed *H. influenzae* to be more abundant in participants with asthma in Uganda and Ecuador, compared to Brazil or NZ.

When repeated for non-asthmatic participants, between-country differences were again identified despite the smaller sample size (n=124) (**Supplementary Figure 2A**). These differences were most pronounced between Ecuador and NZ (pseudo-F=5.82, *P*<0.001), and least evident between Brazil and Uganda (pseudo-F=1.44, *P*=0.15, **Supplementary Table 3**). The patterns of α-diversity and total bacterial load in non-asthmatics replicated those with asthma, with Uganda again having the highest α-diversity and bacterial abundance (**Supplementary Figure 2B-G**).

### Sputum microbiota characteristics differ by asthma status

Microbiota composition differed significantly between participants with asthma and without (R^2^=0.43%, *P*=0.012, **Figure 2A** and **Supplementary Table 3**). Investigation of compositional differences identified no significant association between asthma status and α-diversity measures (**Figure 2B-D**), total bacterial load, or *H. influenzae* and *M. catarrhalis* abundance (**Figure 2E-G**). However, the relative abundance of *Streptococcus* and *Granulicatella* were found to be higher in participants with asthma, while five genera (*Campylobacter, Peptostreptococcus, Leptotrichia, Fusobacterium* and *Mogibacterium*) were less abundant (**Figure 2H, Supplementary Figure 3**).

**Figure 2:**
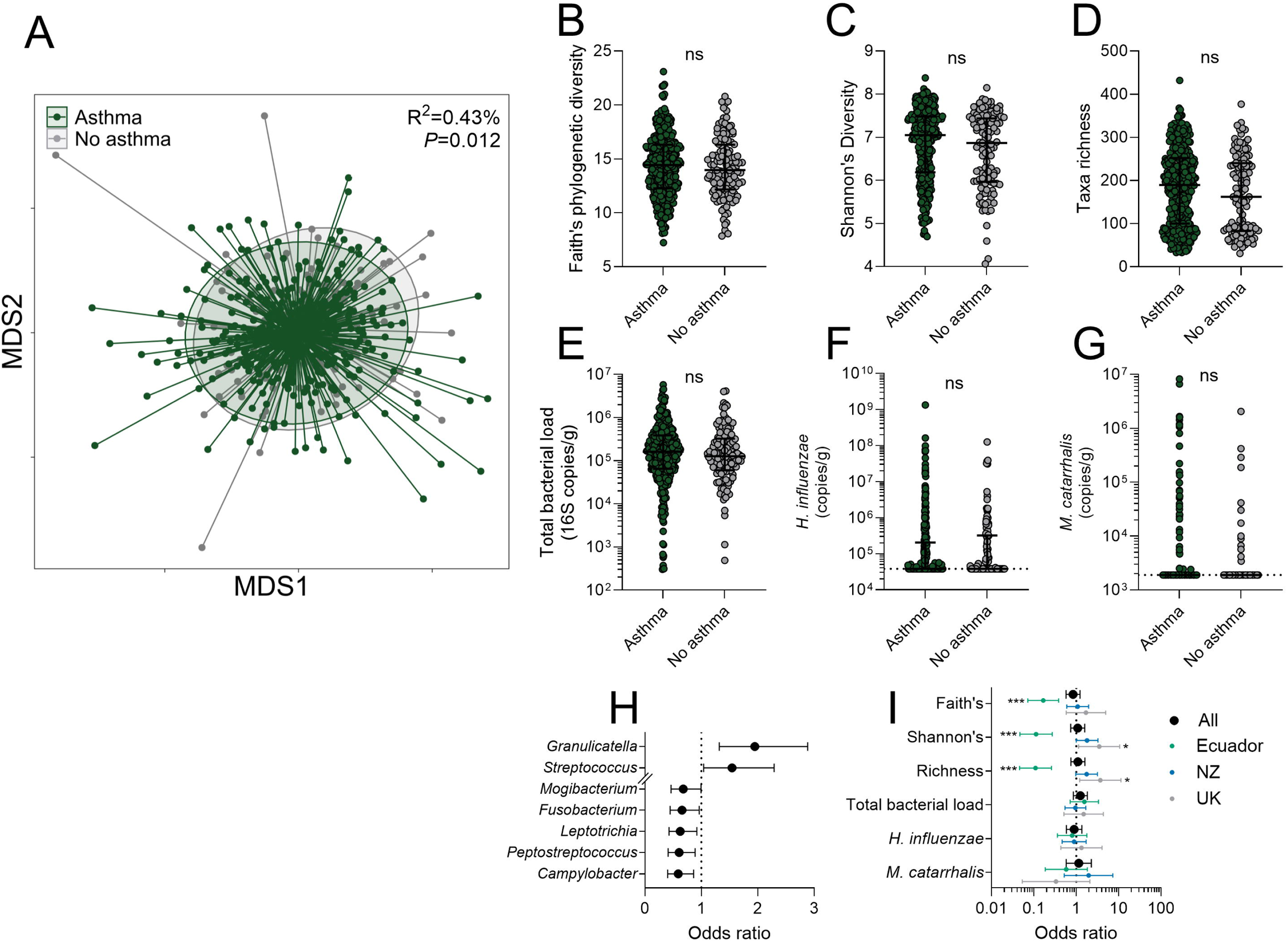
Sputum microbiota differs by asthma status. A) Nonmetric multidimensional scaling (NMDS) plot of Bray-Curtis dissimilarity. B) Faith’s phylogenetic diversity. C) Shannon’s diversity. D) Taxa richness. E) Total bacterial load derived from qPCR. F) *Haemophilus influenzae* abundance derived from qPCR. G) *Moraxella catarrhalis* abundance derived from qPCR. H) Forest plot showing taxa that differed significantly by asthma status; left = lower in asthma. I) Forest plot showing association between asthma and α-diversity (Faith’s, Shannon’s, taxa richness) and qPCR derived bacterial load (total, *H. influenzae* and *M. catarrhalis*); left = lower in asthma. Statistics: A) Permutational multivariate analysis of variance including variables: asthma, country, age and sex; B-I) Ordinal logistic regression including variables: asthma, country, age and sex; *** adjusted *P*<0.001, ** adjusted *P*<0.01, * adjusted *P*<0.05.

Within-country analysis was then performed, with the exclusion of Brazil and Uganda due to low numbers of non-asthmatics. Microbiota composition differed between participants with asthma and without in Ecuador (R^2^=2.74%, *P*<0.001) and NZ (R^2^=1.98%, *P*=0.002), but not the UK (R^2^=1.49%, *P*=0.84). For Ecuador, this was characterised by lower α-diversity in participants with asthma (**Figure 2I**), while in the UK, taxa richness and Shannon’s diversity were higher (**Figure 2I**). Total bacterial load and abundance of *H. influenzae* or *M. catarrhalis* were not associated with asthma status in any country (**Figure 2I**).

### Asthma inflammatory phenotypes are associated with sputum microbiota

The relationship between microbiota composition and inflammatory phenotypes (EA, NA, PGA, and MGA; **Figure 3A-B**) was assessed in participants with asthma. Microbiota composition differed significantly between phenotypes (R^2^=1.71%, *P*<0.001, **Figure 3C, Supplementary Table 4**). Pairwise analysis revealed these differences to be most pronounced between NA and PGA (pseudo-F=3.56, *P*<0.001) and between NA and EA (pseudo-F=3.50, *P*<0.001), with EA also significantly different to PGA (pseudo-F=2.25, *P*=0.015, **Table E4**). Total bacterial load and α-diversity did not differ between inflammatory phenotypes, however *H. influenzae* was significantly higher in NA compared to EA (aOR=2.37 [95% CI: 1.18-4.78], *P=*0.016) or PGA (aOR=3.31 [95% CI: 1.67-6.54], *P*<0.001, **Supplementary Figure 4**). Analysis of differences in taxon distribution using LEfSe identified an unassigned Lachnospiraceae taxon as enriched in PGA (LDA score=3.5; *P*=0.032), but no other significant associations.

**Figure 3:**
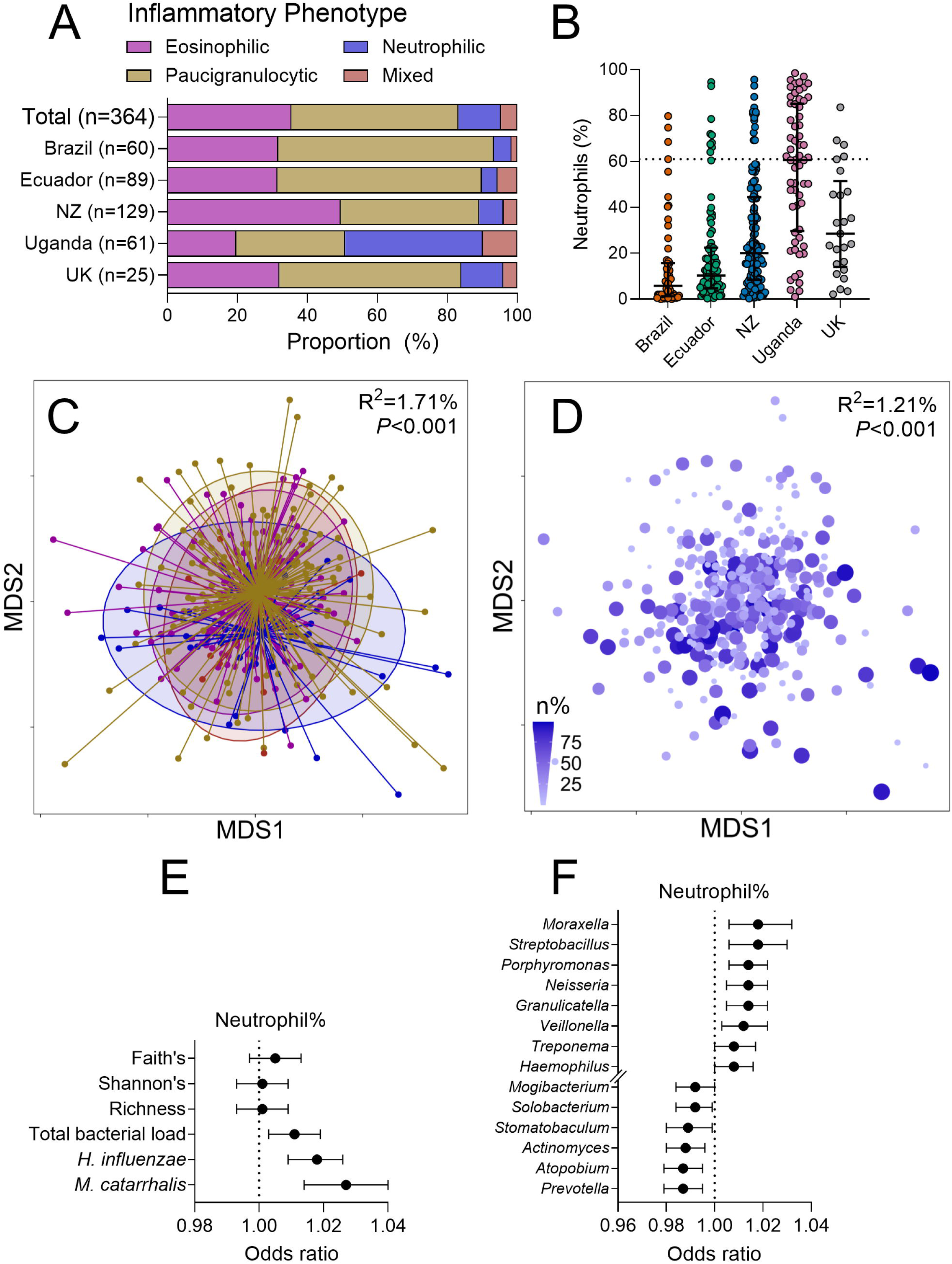
Sputum microbiota differs by inflammatory phenotype in asthma. A) Distribution of inflammatory phenotypes by country. B) Distribution of sputum neutrophil% by country. C) Nonmetric multidimensional scaling (NMDS) plot of Bray-Curtis dissimilarity by inflammatory phenotype where eosinophilic = purple, paucigranulocytic = gold, neutrophilic = blue, mixed = red. D) NMDS plot showing dispersion by neutrophil percentage (n%). E) Forest plot showing neutrophil% associated with α-diversity (Faith’s, Shannon’s, richness) and qPCR derived bacterial load (total, *H. influenzae* and *M. catarrhalis*). F) Forest plot showing taxa that differed significantly by neutrophil%. Statistics: C) Permutational multivariate analysis of variance including variables: inflammatory phenotype, country, age and sex; D) Permutational multivariate analysis of variance including variables: neutrophil%, eosinophil%, country, age and sex; E, F) Ordinal logistic regression including variables: neutrophil%, eosinophil%, country, age and sex; *** adjusted *P*<0.001, ** adjusted *P*<0.01, * adjusted *P*<0.05.

When each inflammatory phenotype was compared to the non-asthmatic group, significant divergence was observed for NA (pseudo-F=3.59, *P*<0.001) and EA (pseudo-F=2.93, *P*=0.003), but not for PGA (pseudo-F=1.40, *P*=0.14) or MGA (pseudo-F=1.06, *P*=0.40) (**Supplementary Table 4**).

To further explore associations between airway inflammation and sputum microbiota composition, analyses were performed using both neutrophil% and eosinophil% as separate continuous measures within the same model. Both neutrophil% and eosinophil% were significantly associated with microbiota composition, with this relationship being more pronounced for neutrophil% (neutrophil%: R^2^=1.21%, *P*<0.001, **Figure 3D**; eosinophil%: R^2^=0.49%, *P*=0.033, **Supplementary Figure 5A**; **Table 2**).

**Table 2:** Permutational multivariate analysis of variance (PERMANOVA) output assessing the independent effect of sputum neutrophil and eosinophil percentage in participants with asthma.

|  | **R<sup>2</sup> (%)** | **Pseudo-F** | **P value** | **Significance** |
| --- | --- | --- | --- | --- |
| **Univariate** |  |  |  |  |
| Neutrophil % | 1.21 | 4.43 | <0.001 | *** |
| Eosinophil % | 0.45 | 1.62 | 0.087 |  |
| **Multivariate** |  |  |  |  |
| Neutrophil % | 1.21 | 4.79 | <0.001 | *** |
| Eosinophil % | 0.49 | 1.94 | 0.033 | * |
| Age | 0.90 | 3.56 | <0.001 | *** |
| Sex | 0.46 | 1.80 | 0.051 |  |
| Country | 7.65 | 7.58 | <0.001 | *** |

Significance codes: \*\*\* $P < 0.001$ , \* $P < 0.05$ .

No α-diversity metric was associated with either neutrophil% (**Figure 3E**) or eosinophil% (**Supplementary Figure 5C**). However, neutrophil% (**Figure 3E**), but not eosinophil% (**Supplementary Figure 5C**), was associated with a higher bacterial load (*P=*0.0089). Across all taxa, neutrophil% was positively associated with eight individual taxa and negatively associated with six (**Figure 3F**). These included positive associations with relative abundance of *Haemophilus* and *Moraxella*, which was also reflected by positive association with absolute abundance of *H. influenzae* and *M. catarrhalis* (**Figure 3E**). Analysis based on eosinophil% identified positive associations with *Streptococcus*, *Streptobacillus*, and *Actinobacillus*, and negative associations with *Prevotella* and *Leptotrichia* (**Supplementary Figure 5D**).

### Inflammation-microbiota relationships are country-specific

We next assessed whether the associations between markers of airway inflammation and sputum microbiota characteristics were country specific. Due to the smaller number of participants from each country, this analysis was performed using inflammatory cell percentage, rather than phenotype groups, with the UK subgroup excluded. While the analysis across the whole study showed that neutrophilia was positively associated with total bacterial load and microbiota similarity, associations within each country were more varied. In NZ, neutrophil% was significantly associated with microbiota similarity (R^2^=2.51%, *P*<0.001, **Supplementary Table 5**). Similar relationships in Brazil and Uganda did not achieve statistical significance and no association was observed for Ecuador (**Supplementary Table 5**).

All three α-diversity metrics were inversely associated with neutrophil% in Brazil, but no other country (**Figure 4A, Supplementary Figure 6B-D**). Associations between neutrophil% and bacterial taxa differed between countries. Notably, *Neisseria* was positively associated with neutrophil% in NZ and Uganda, while *Granulicatella* was positively associated with neutrophil% in Brazil and Ecuador (**Figure 4B-E**).

**Figure 4:**
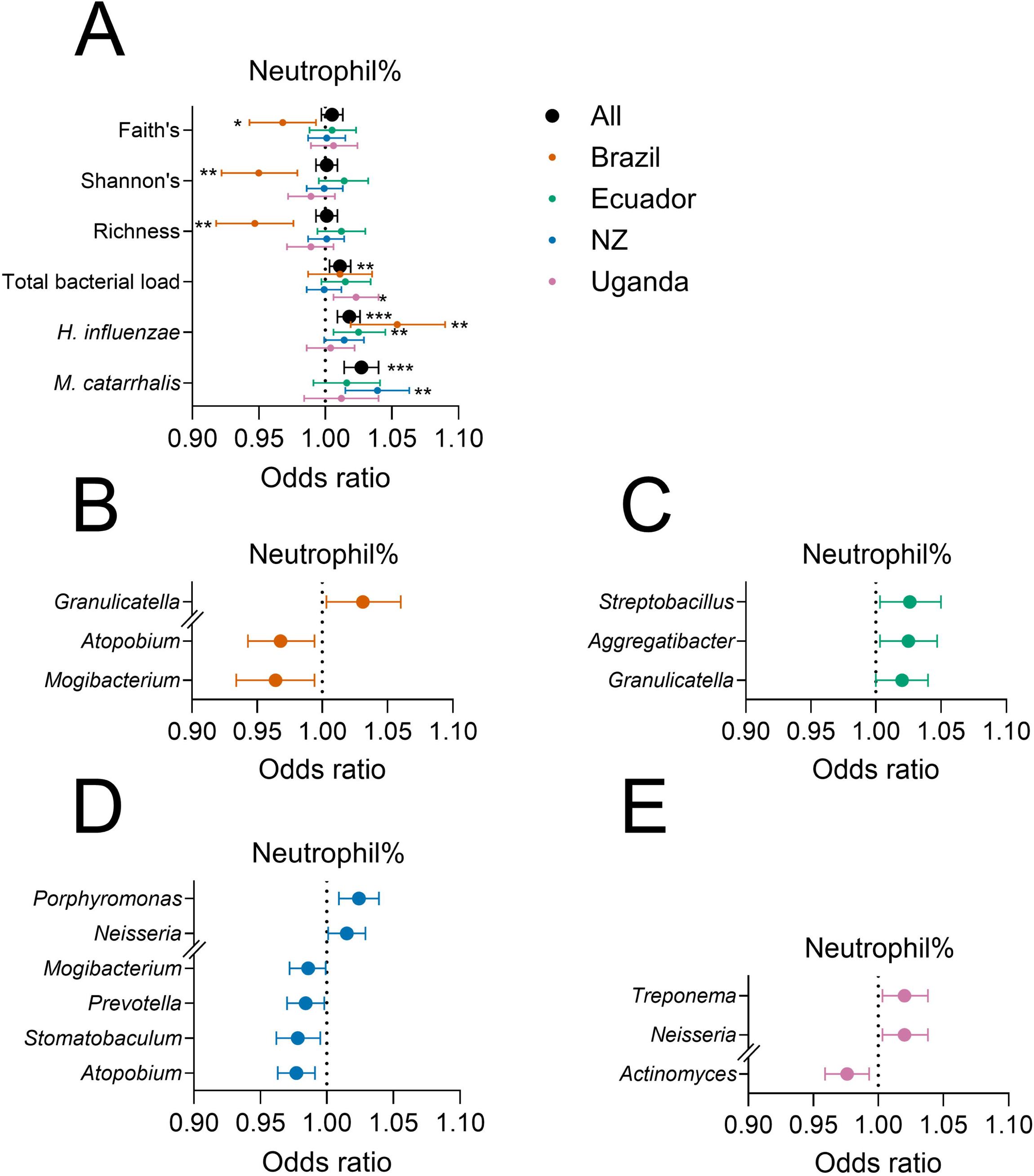
Within country association between sputum microbiota and neutrophil%. A) Forest plot showing neutrophil% associated with α-diversity (Faith’s, Shannon’s, richness) and qPCR derived bacterial load (total, *H. influenzae* and *M. catarrhalis*) in whole cohort (black), and within Brazil (orange), Ecuador (green), New Zealand (blue), and Uganda (pink). B) Forest plot showing taxa that differed by neutrophil% in Brazil. C) Forest plot showing taxa that differed by neutrophil% in Ecuador. D) Forest plot showing taxa that differed by neutrophil% in New Zealand. E) Forest plot showing taxa that differed by neutrophil% in Uganda. Statistics: Ordinal logistic regression including variables: neutrophil%, eosinophil%, age and sex; *** adjusted *P*<0.001, ** adjusted *P*<0.01, * adjusted *P*<0.05. *M. catarrhalis* analysis was not performed for Brazil due to low detection frequency (3 out of 60 participants).

Total bacterial load, and abundance of *H. influenzae* and *M. catarrhalis* varied within countries, with total bacterial load positively associated with neutrophil% in Uganda, Brazil, and Ecuador, but only reaching significance for Uganda (*P=*0.032, **Figure 4A, Supplementary Figure 6A**). *H. influenzae* load was positively associated with neutrophil% in Brazil (*P=*0.002), Ecuador (*P=*0.008), and weakly associated in NZ (*P=*0.065), but not associated in Uganda (*P=*0.67). *M. catarrhalis* was positively associated with neutrophil% in NZ (*P=*0.002), but in no other country (**Figure 4A, Supplementary Figure 6E-F**).

Eosinophil% was not significantly associated with microbiota composition in any country, although a borderline statistically significant association was observed for Brazil (R^2^=3.08%, *P*=0.051, **Supplementary Table 5**). Eosinophil% was inversely associated with Shannon’s diversity and richness in Uganda, and a trend towards a positive association was observed for NZ (**Supplementary Figure 7A**). Total bacterial load and absolute abundance of *H. influenzae* and *M. catarrhalis* was not associated with eosinophil% in any country (**Supplementary Figure 6A**). Of the eight taxa associated with eosinophil% (**Supplementary Figure 7B-E**), *Streptococcus* was notable in being positively associated with eosinophil% in both Brazil and Uganda.

## DISCUSSION

We previously reported that sputum inflammatory phenotypes in young people with asthma differed between the assessed countries, with higher levels of neutrophilic inflammation in Uganda and higher levels of eosinophilic inflammation in NZ [9]. Here, we build on this prior study to report four principal findings in relation to global asthma heterogeneity and its relationship with microbiology: 1) airway microbiota characteristics differ between countries; 2) and between asthmatic and non-asthmatic participants; 3) microbiota characteristics are strongly associated with levels of neutrophilic inflammation; and 4) microbiota characteristics are associated with eosinophilic inflammation, but this relationship is less pronounced than for neutrophilia.

We identified significant differences in airway microbiota characteristics between participant groups from each of the five countries. For example, the most pronounced differences in microbiota composition were observed between NZ and Uganda, while Brazil and Ecuador were most similar. Airway microbiota in both asthmatic and non-asthmatic participants from Uganda was characterised by a higher diversity and bacterial load, and a higher abundance of *H. influenzae*. Such cohort-specific traits could reflect a range of factors including differences in microbial exposure or differences in environmental, lifestyle, and socioeconomic factors that influence the ability of microbes to colonise the airways.

Previous efforts to explore differences in the airway microbiota between asthma and non-asthma have focused largely on older populations in high income settings [3, 24–29]. We identified a significant but modest difference in microbiota composition between asthmatic and non-asthmatic participants across the five countries assessed. While this was not associated with differences in diversity or bacterial load, several common oropharyngeal organisms were differentially associated with asthma, independent of age, sex, and country. Associations between asthma and airway microbes in the literature vary, but there are some consistencies with the findings reported here. Namely, the higher levels of *Granulicatella* and *Streptococcus* we observed in participants with asthma are supported by previous investigations by Durack *et al*. [3] and Goleva *et al*., [27] respectively. In a separate study by Durack *et al*., the authors also reported lower levels of *Leptotrichia*, and *Peptostreptococcus* in participants with asthma compared to without, again consistent with our findings [25]. As yet, the basis for these associations are unclear.

Despite between-country differences in both airway microbiota characteristics and inflammatory phenotypes, a consistent association between both was observed after adjustment for age, sex, and country. Overall, neutrophilic inflammation was associated with microbiota composition, showing a positive relationship with total bacterial load, abundance of *H. influenzae*, *M. catarrhalis*, as well as other potentially pathogenic taxa, such as *Neisseria* and *Veillonella*.

The association between airway neutrophilia and pathogen or pathobiont abundance in asthma is supported by findings of studies in adults in HICs [4, 5, 30–33], and may reflect specific bidirectional processes. For example, both increased and altered neutrophil function in asthma can facilitate airway colonisation by opportunistic organisms, while presence of pathobionts may promote neutrophil recruitment [34]. Although other studies have reported an inverse association between neutrophils and microbiota diversity, this was not observed here, potentially reflecting the age of participants. In particular, more established neutrophilic disease in adult asthma, and associated therapies, is likely to exert a greater selective pressure on microbiota composition compared to younger individuals with milder disease.

In contrast to neutrophil levels, the association with eosinophilic inflammation with airway microbiota characteristics was less pronounced. Specifically, while microbiota composition of EA was significantly different to PGA and no asthma, the microbiota association with neutrophil% was 2.46 times stronger compared to eosinophil% (1.21% compared to 0.49%). Further, eosinophil% was not associated with bacterial load, or the detection of potential airway pathogens. However, eosinophil% was positively associated with *Streptococcus* abundance, the most prevalent and abundant taxa. These findings are in keeping with previous studies that have reported various strengths of association between eosinophilic inflammation and the airway microbiota [2, 3, 24, 32, 33, 35], from none or minimal [2, 32], to moderate or strong [3, 24].

Despite small sample sizes, within-country analysis did identify region-specific associations. A greater divergence in microbiota composition between participants with or without asthma was observed in Ecuador, characterised by lower microbiota diversity in participants with asthma. Similarly, in those with asthma, microbiota diversity was inversely associated with neutrophil% in Brazil, while pathobiont abundance was positively associated with neutrophil% in Brazil, Ecuador, and NZ. The lack of an association between neutrophil% and *H. influenzae* and *M. catarrhalis* levels in the Ugandan cohort is notable, given that individuals from Uganda exhibited higher neutrophil%, total bacteria, and these specific pathobionts. While a positive association between neutrophil% and total bacteria in Uganda was observed, the lack of an association with *H. influenzae* and *M. catarrhalis* may reflect the contribution of other pathobiont species. Indeed, *Neisseria* and *Treponema* spp. were positively associated with neutrophil% in Uganda, with both genera including species with pathogenic potential [36, 37].

Whether the region-specific associations identified here represent specific mechanistic processes is unclear. However, the combined influence of differences in inflammatory phenotype prevalence and environmental, lifestyle and socioeconomic factors could be significant. Regardless, these differences highlight the need for regional consideration when studying associations between asthma phenotypes and airway microbiota. These relationships may also have implications for asthma management strategies in different geographical regions. For example, the efficacy of maintenance macrolide therapy has been shown to be predicted by abundance of *H. influenzae* in adults with persistent uncontrolled asthma [7].

Important strengths of this study were the international representation of asthma and the detailed assessment of inflammatory phenotypes and airway microbiology. To our knowledge, it represents the most diverse single asthma airway microbiota study to date. With such a diverse cohort, careful protocol alignment for sample collection and processing was necessary. All sputum DNA extraction, sequencing, and bioinformatic processing was performed at a single site, with careful inclusion of extraction and sequencing controls to assess and account for batch effects. Further, all analyses were adjusted for age and sex to account for between-group variance. We also undertook multiple approaches to explore the inflammation-microbiota relationships, including the conventional four group assessment, as well as including both neutrophil% and eosinophil% in a single model to enable mutual adjustment.

There are several important considerations when interpreting our findings. While the WASP-biome cohort was representative of the original WASP study, global variance of asthma phenotypes is broad [38] and the generalisability of our findings requires further investigation. Asthma cases were identified from responses to the ISAAC questionnaire, which assesses factors such as wheezing, whistling in the chest, or asthma medication use. While it does not include objective tests (such as bronchodilator response or airway hyperreactivity testing), this validated questionnaire is particularly applicable in LMICs, where clinical testing is difficult and rates of physician-diagnosed asthma are low [39]. Finally, 16S rRNA amplicon sequencing used to assess the microbiota has limited taxonomic resolution beyond genus-level, and does not capture fungi or viruses, which may be associated with asthma heterogeneity.

In conclusion, we report variance in associations between airway microbiology and inflammatory phenotypes between countries, highlighting the need for further global studies of asthma pathophysiology, with potential implications for diagnosis and selection of therapies. We also report conserved associations between the airway microbiota and inflammatory phenotypes across HICs and LMICs, particularly in neutrophilic asthma. With growing efforts to personalise asthma treatment and management strategies, there is now a clear opportunity to further examine how immune-microbiota interactions may guide such approaches.

## Data Availability

Reads have been deposited in the European Bioinformatics Institute European Nucleotide Archive (PRJEB77703).

## Contributions

Conceptualisation: NP, CRB, HM, MB, PJC, and JD; Data Curation: SLT, LP, CRB, JB, HM, CAF, AYO, MC, HA, IN, PT, SRi, MB, PJC, AAC, JD, NP; Formal Analysis: SLT; Funding Acquisition: NP, LP, JD, SLT; Investigation: SLT, LE, SKM; Methodology: SLT, CRB, JD, GBR; Resources: LP, CER; Validation: SLT, LE, SKS; Visualisation: SLT; Writing – Original Draft Preparation: SLT, GBR, CRB, JD; Writing – Review & Editing: SLT, LE, SKM, LP, CRB, HM, CAF, AYO, MC, JB, HA, IN, PT, SRi, CER, KvV, SR, MB, PJC, AAC, NP, GBR, JD.

## The WASP Study group

<u>United Kingdom, London:</u> Neil Pearce*, Lucy Pembrey*, Steven Robertson*, Karin van Veldhoven*, Charlotte E Rutter*, Sinead Langan, Sarah Thorne, Donna Davoren

<u>United Kingdom, Bristol:</u> John Henderson, Susan Ring*, Elizabeth Brierley, Sophie Fitzgibbon, Simon Scoltock, Amanda Hill

<u>Brazil, Leading Group:</u> Alvaro Cruz*, Camila Figueiredo*, Mauricio Barreto*

<u>Brazil, ProAR collaborators/associates:</u> Cinthia Vila Nova Santana, Gabriela Pimentel Pinheiro, Gilvaneide Lima, Valmar Bião Lima, Jamille Fernandes

<u>Brazil, Lab students/associates:</u> Tamires Cana Brasil Carneiro, Candace Andrade, Gerson Queiroz, Anaque Pires, Milca Silva, Jéssica Cerqueira

<u>Ecuador:</u> Philip Cooper*, Martha Chico, Cristina Ardura-Garcia, Araceli Falcones, Aida Y Oviedo, Andrea Zambrano

<u>New Zealand:</u> Jeroen Douwes*, Collin Brooks*, Hajar Ali*, Jeroen Burmanje*

<u>Uganda:</u> Harriet Mpairwe*, Irene Nambuya, Pius Tumwesige, Milly Namutebi, Marble Nnaluwooza, Mike Mukasa

*Writing Group for this paper

## Acknowledgements

The World ASthma Phenotypes (WASP) collaboration is based on the Asthma Phenotypes study which is funded by the European Research Council under the European Union’s Seventh Framework Programme (FP7/2007–2013)/ERC grant agreements 668954 and 101020088. The UK Medical Research Council and Wellcome (21706/Z/19/Z) and the University of Bristol provide core support for ALSPAC. The study was supported by the Marsden Fund (20-MAU-071) and the New Zealand Health Research Council (15/311, 14/474). SLT is supported by a National Health and Medical Research Council (NHMRC) Investigator Grant (2008625) and CRB is supported by an NZ HRC Sir Charles Hercus Fellowship. We would like to thank the participants of this study in all five WASP centres.

