## Supplementary material for "Airway microbiota in young people across four continents differ by country, asthma status and inflammatory phenotype": Online Supplement

**Ethics approval**

Ethics approval for the study has been obtained from the London School of Hygiene & Tropical Medicine (LSHTM) ethics committee (ref: 9776) and in all five study centres. Informed consent was obtained from all participants or their parents/carers before taking part.

Avon Longitudinal Study of Parents and Children (ALSPAC): Ethical approval for the UK-arm of the study was obtained from the ALSPAC Ethics and Law Committee, and the Local Research Ethics Committees. Consent for biological samples has been collected in accordance with the Human Tissue Act (2004). REDcap was used to collect the ALSPAC data (http://projectredcap.org/resources/citations/). Informed consent for the use of data collected via questionnaires and clinics was obtained from participants following the recommendations of the ALSPAC Ethics and Law Committee at the time. The ALSPAC study website contains details of all the data that is available through a fully searchable data dictionary and variable search tool (http://www.bristol.ac.uk/alspac/researchers/our-data/).

**Recruitment**

Participants with and without asthma were recruited from the same sources, which varied between sites. For the UK, both asthmatic and non-asthmatic participants were recruited through the ongoing ALSPAC study. For Brazil, Ecuador, and New Zealand, recruitment of participants with and without asthma was through both existing cohort studies[1, 2] and additional community recruitment (usually from surveys in schools). In Uganda, participants were recruited from a larger case-control study of asthma identified through a cross-sectional survey in schools [3].

**Sputum induction**

Sputum induction was performed when participants were stable. Aerosolised hypertonic saline (4.5% w/v) was produced using an ultrasonic nebuliser (DeVilbiss Ultraneb 2000, Langen, Germany) and administered orally through a mouthpiece (Hans-Rudolph Inc, Kansas City, USA) for increasing intervals from 0.5-4 minutes, to a total of 15.5 minutes. Spirometry was conducted between intervals, and salbutamol was administered if forced expiratory volume in 1 second (FEV_1_) dropped to ≤75%-predicted. Participants were subsequently encouraged to produce sputum in a sterile plastic container. Sputum was separated into aliquots, with one stored unprocessed at -80°C for microbiota analysis, and others processed for inflammatory phenotyping.

**Sputum inflammatory phenotyping**

Inflammatory phenotyping was performed as described previously [4], using well-characterised protocols [5]. Sputum plugs were dispersed using dithiothreitol (Sputasol, Oxoid Ltd, Hampshire, England), filtered through a 60 µm filter (Millipore, County Cork, Ireland), centrifuged at 400 × g, and resuspended in phosphate buffered saline (PBS). The resulting cell suspension used to prepare cytospin slides stained using a Diff-QuikVR fixative and stain set (Dade Behring, Deerfield, IL). Sputum slides were read in Wellington, New Zealand, with the exception of the slides produced in Brazil (which could not be shipped overseas due to ethical restrictions): these were therefore read in Brazil, with a sample of slides being remotely checked (using microscopy images) by the group in Wellington.

**Sputum DNA extraction**

All samples underwent DNA extraction at a single facility, with extraction batching including samples from at least three countries to minimise batch effects. For each sample, 100 mg of unprocessed induced sputum was suspended in 300 µL of PBS, vortexed for 10 seconds, and placed on ice for 2 min. Samples were pelleted by centrifugation at 13,000 × g for 10 min. Supernatant was removed and the pellet was resuspended in 300 μL of Tris-EDTA solution (10 mM Tris-HCl, 1 mM EDTA; pH 8.0; Ambion, ThermoFisher Scientific, Victoria, Australia), 200 mg of silica: zirconium beads (1:1 of 0.1 mm and 1.0 mm; Biospec Products, Inc., OK, USA), and a single chrome bead (3.2 mm, Biospec Products, Inc., OK, USA) added. Samples underwent bead-beating at 6.5 m/s for 60 sec in a FastPrep®-24 Instrument (MP Biomedicals, CA, USA). Homogenised samples were heated to 95°C for 1 min, before being cooled on ice for 1 min. Lysozyme (ROCHE, ThermoFisher Scientific, Victoria, Australia) and lysostaphin (Sigma-Aldrich, MO, USA) were then added to a final concentration of 2 mg/mL and 0.1 mg/mL, respectively, and samples incubated at 37°C for 1 hr. Proteinase K (Fermentas, ThermoFisher Scientific, Victoria, Australia) and sodium dodecyl sulphate (Sigma-Aldrich, MO, USA) were then added to a final concentration of 1.2 mg/mL and 1.5 %, w/v, respectively. Following incubation at 30 min at 56°C, 40 μL of 5 M sodium chloride and 450 μL of phenol:chloroform:isoamyl alcohol (25:24:1; saline buffered at pH8.0; Sigma-Aldrich, MO, USA) were added and samples vortexed for 30 sec. The aqueous-organic layers were separated by centrifugation at 13,000 × g for 10 min at 4°C and 400 μL of the aqueous layer was transferred to a new microfuge tube. DNA was recovered using an EZ-10 Spin column in accordance with manufacturer’s instructions (Bio Basic, Inc., Ontario, Canada), following precipitation by the addition of 10 M ammonium acetate and 99% ethanol (Sigma Aldrich, MO, USA) in a 1:10 and 1:1 ratio with sample volume, respectively. DNA was eluted in 100 μL UltraPure DNase/RNase-free distilled water (Gibco, ThermoFisher Scientific, Victoria, Australia) and stored at -80°C prior to analysis.

**Quantitative PCR**

Abundance of all bacteria (16S), *Haemophilus influenzae* (*smpB*)*, Moraxella catarrhalis* (*copB*) were measured by quantitative PCR (qPCR) as described previously [6, 7], using the primers and cycling conditions in **Table E1**. All assays were performed on the QuantStudio 6 Flex System (Thermo Fisher Scientific, Vic, Australia). Reactions were performed in triplicate and averages taken. Gene copy numbers was calculated against a standard curve of a known bacterial concentration and normalised per g of sputum.

**16S rRNA gene amplicon sequencing**

The V1-3 hypervariable region of the bacterial 16S rRNA gene was amplified from sputum DNA using modified primers 27F (5'-TCGTCGGCAGCGTCAGATGTGTATAAGAGACAGAGRGTTTGATCMTGGCTCAG-3') and 519R (5'-GTCTCGTGGGCTCGGAGATGTGTATAAGAGACAGGTNTTACNGCGGCKGCTG-3'), with Illumina adapter overhang sequences as indicated by underline. Amplicons were generated, cleaned, indexed and sequenced according to the Illumina MiSeq 16S Metagenomic Sequencing Library Preparation protocol with certain modifications. Briefly, an initial PCR reaction contained at least 12.5 ng of DNA, 5 μL of forward primer (1 μM), 5 μL of reverse primer (1 μM) and 12.5 μL of 2 × KAPA HiFi Hotstart ReadyMix (KAPA Biosystems, Wilmington, MA, USA) in a total volume of 25 μL. The PCR reaction was performed on a Veriti 96-well Thermal Cycler (Life Technologies) using the following program: 95 °C for 3 min, followed by 25 cycles of 95 °C for 30 sec, 55 °C for 30 sec and 72 °C for 30 sec and a final extension step at 72 °C for 5 min. Samples were multiplexed using a dual-index approach with the Nextera XT Index kit (Illumina Inc., San Diego, CA, USA) according to the manufacturer’s instructions. The final library was paired-end sequenced at 2 × 300 bp using a MiSeq Reagent Kit v3 on the Illumina MiSeq platform. Sequencing was performed at the South Australian Genomics Centre.

**Bioinformatic processing**

Demultiplexed sequences were processed using QIIME2 [8] (release 2021.11).Trimmed paired-end reads were merged, chimeric sequences removed, denoised sequencing errors corrected using the DADA2 plugin [9]. Representative sequences were aligned to the SILVA database (v138) at 80% using vsearch and unassigned sequences were filtered out. Remaining unique amplicon sequence variants (ASVs) were classified using the QIIME2 sklearn algorithm to the SILVA database at 99% sequence similarity. ASVs that were amplified in the blank extraction control were examined. Contaminant or spurious taxa, not associated with the human microbiota were filtered out. The median clean and filtered read count was 4398 (IQR: 3086, 5861). Samples were rarefied to 1463 reads, as determined by alpha rarefaction plot. The resulting ASVs were used to calculate α-diversity (Faith’s phylogenetic diversity, Shannon’s diversity, and taxon richness), while the genus-level taxonomic assignment was converted to relative abundances.

**Data analysis and visualisation**

Analysis of dissimilarity in square-root transformed, rarefied, genus-level microbiota composition (β-diversity) was performed by permutational multivariate analysis of variance (PERMANOVA) using Bray–Curtis distances, with the adonis2 function from the ‘*vegan’* R package (v2.5-7) [10], with 9999 permutations. To assess whether unbalanced designs impacted the PERMANOVA findings, assessments were repeated in Primer (v6.1.16; PRIMER-E, Plymouth, United Kingdom) using a Type III model that accounts for unbalanced designs. Differences in α-diversity metrics, bacterial load, and relative abundance of specific taxa were calculated using ordinal logistic regression using SAS Studio (v3.81; SAS Institute Inc., NC, USA). Differences in taxon relative abundance between groups of three or more (countries and inflammatory phenotypes) were performed by linear discriminant analysis (LDA) Effect Size (LEfSe) [11], using one-against-all multi-class analysis, and cut-offs of LDA ≥3 and p<0.05. Findings from LEfSe were confirmed by Wilcoxon signed rank test. Holm–Bonferroni multiple comparison correction was applied when analyses of multiple dependent variables were performed.

Microbiota dissimilarity between individuals was visualised using non-metric multidimensional scaling (nMDS), calculated using the ‘*vegan’* R package. All data were visualised using GraphPad Prism (v10.0.2) except for nMDS plots and taxa bar plots, which were visualised using the ‘*ggplot2*’ R package (v3.3.5).

**Supplementary** **Table 1:** Primers and cycling conditions for quantitative PCR

| Target | Primers (5'-3') | Cycling conditions | Ref |
| --- | --- | --- | --- |
| 16S | F: TCCTACGGGAGGCAGCAGT  R: GGACTACCAGGGTATCTAATCCTGTT | 95 °C for 15 s,  60 °C for 1 min | [12] |
| *smpB* | F: ATTAAATGTTGCATCAACGC  R: GACTTTTGCCCACGCAC  Probe: FAM- ACGRTTTTACCATAGTTGCACTTTCTC- BHQ | 95 °C for 10 s,  63 °C for 1 min | [13] |
| *copB* | F: GTGAGTGCCGCTTTTACAACC  R: TGTATCGCCTGCCAAGACAA | 95 °C for 15 s,  60 °C for 1 min | [14] |

**Supplementary** **Table 2:** Demographic and asthma characteristics of WASP and WASP-biome

|  | **WASP** | **WASP-biome** |
| --- | --- | --- |
| All (n) | 920 | 488 |
| Female, n (%) | 526 (57.2%) | 256 (52.5%) |
| Age (years), median (IQR) | 15.57 (11.76-17.93) | 14.11 (11.41-17.35) |
| Asthma (n) | 658 | 364 |
| Female, n (%) | 387 (58.8%) | 198 (54.4%) |
| Age (years), median (IQR) | 15.34 (11.94-17.93) | 14.08 (11.17-17.15) |
| BMI (kg/m^2^), median (IQR) | 20.80 (18.12-24.22) (n=655) | 20.44 (17.87-23.49) |
| ACQ7 score, median (IQR) | 0.50 (0.00-1.17) (n=631) | 0.50 (0.00-1.17) |
| ACQ7 level, n (%) | (n=632) |  |
| Well controlled | 503 (79.6%) | 290 (81.5%) |
| Uncontrolled | 129 (20.4%) | 66 (18.5%) |
| ICS use past 12 months, n (%) | 255 (43.1%) (n=592) | 118 (36.9%) |
| BA use past 12 months, n (%) | 402 (67.3%) (n=597) | 202 (62.5%) |
| FEV_1_ (% predicted), mean (STD) | 94.10 (12.46) (n=628) | 95.00 (12.27) |
| FVC (% predicted), mean (STD) | 98.48 (12.98) (n=628) | 99.07 (12.30) |
| Atopy, n (%) | 432 (67.2) (n=643) | 225 (63.0%) |
| Inflammatory phenotype, n (%) |  |  |
| Eosinophilic | 239 (36.3%) | 131 (36.0%) |
| Neutrophilic | 68 (10.3%) | 43 (11.8%) |
| Paucigranulocytic | 328 (49.9%) | 172 (47.3%) |
| Mixed granulocytic | 23 (3.5%) | 18 (4.9%) |
| Sputum neutrophils (%), median (IQR) | 15.85 (5.50-39.86) | 17.71 (6.09-47.50) |
| Sputum eosinophils (%), median (IQR) | 1.45 (0.00-6.48) | 1.50 (0.25-6.28) |

IQR: Interquartile range, STD: Standard deviation, BMI: Body mass index, ACQ7: Asthma control questionnaire, ICS: Inhaled corticosteroids, BA: Beta-agonist, FEV_1_: Forced expiratory volume in 1 second, FVC: Forced vital capacity

**Supplementary** **Table 3**: Permutational multivariate analysis of variance (PERMANOVA) output assessing the effect of country on microbiota composition in asthma

|  | **R^2^ (%)** | **Pseudo-F** | ***P* value** | **Significance** |
| --- | --- | --- | --- | --- |
| **Univariate: Asthma (n=364)** | | | | |
| Age | 2.03 | 7.51 | <0.001 | *** |
| Sex | 0.63 | 2.30 | 0.010 | ** |
| Country | 7.75 | 7.54 | <0.001 | *** |
| **Multivariate: Asthma (n=364)** | | | | |
| Age | 0.96 | 3.77 | <0.001 | *** |
| Sex | 0.43 | 1.67 | 0.069 |  |
| Country | 7.75 | 7.61 | <0.001 | *** |
| Brazil vs Ecuador | 1.88 | 2.81 | <0.001 | *** |
| Brazil vs NZ | 4.55 | 8.90 | <0.001 | *** |
| Brazil vs Uganda | 4.87 | 6.09 | <0.001 | *** |
| Brazil vs UK | 4.37 | 3.79 | <0.001 | *** |
| Ecuador vs NZ | 4.75 | 10.80 | <0.001 | *** |
| Ecuador vs Uganda | 6.00 | 9.46 | <0.001 | *** |
| Ecuador vs UK | 4.52 | 5.30 | <0.001 | *** |
| NZ vs Uganda | 5.78 | 11.54 | <0.001 | *** |
| NZ vs UK | 4.19 | 6.65 | <0.001 | *** |
| Uganda vs UK | 5.56 | 4.94 | <0.001 | *** |
| **Multivariate: Non-asthma (n=124)** | | | | |
| Age | 2.21 | 2.96 | 0.015 | * |
| Sex | 1.09 | 1.46 | 0.12 |  |
| Country | 9.29 | 3.11 | <0.001 | *** |
| Brazil vs Ecuador | 5.20 | 1.93 | 0.035 | * |
| Brazil vs NZ | 3.71 | 2.58 | 0.007 | ** |
| Brazil vs Uganda | 9.42 | 1.39 | 0.136 |  |
| Brazil vs UK | 9.78 | 2.71 | 0.005 | ** |
| Ecuador vs NZ | 6.16 | 5.82 | <0.001 | *** |
| Ecuador vs Uganda | 4.63 | 1.74 | 0.080 |  |
| Ecuador vs UK | 10.37 | 5.61 | <0.001 | *** |
| NZ vs Uganda | 2.10 | 1.44 | 0.15 |  |
| NZ vs UK | 2.59 | 2.10 | 0.024 | * |
| Uganda vs UK | 7.08 | 1.99 | 0.030 | * |
| **Multivariate: All (n=488)** | | | | |
| Age | 1.22 | 6.43 | <0.001 | *** |
| Sex | 0.35 | 1.86 | 0.038 | * |
| Asthma | 0.43 | 2.27 | 0.012 | * |
| Country | 6.74 | 8.86 | <0.001 | *** |

Significance codes: *** *P*<0.001, ** *P*<0.01, * *P*<0.05.

**Supplementary** **Table 4**: Permutational multivariate analysis of variance (PERMANOVA) output assessing the effect of inflammatory phenotypes on microbiota composition

|  | **R^2^ (%)** | **Pseudo-F** | ***P* value** | **Significance** |
| --- | --- | --- | --- | --- |
| **Multivariate: Asthma (n=364)** |  |  |  |  |
| Inflammatory phenotype | 1.71 | 2.26 | <0.001 | *** |
| Eosinophilic vs Paucigranulocytic | 0.69 | 2.25 | 0.015 | * |
| Eosinophilic vs Neutrophilic | 1.90 | 3.50 | <0.001 | *** |
| Eosinophilic vs Mixed | 0.65 | 1.04 | 0.40 |  |
| Paucigranulocytic vs Neutrophilic | 1.52 | 3.56 | <0.001 | *** |
| Paucigranulocytic vs Mixed | 0.49 | 1.00 | 0.43 |  |
| Neutrophilic vs Mixed | 0.87 | 0.56 | 0.87 |  |
| Age | 0.92 | 3.64 | <0.001 | *** |
| Sex | 0.45 | 1.80 | 0.042 | * |
| Country | 7.56 | 7.49 | <0.001 | *** |
| **Multivariate: All (n=488)** | | | | |
| Inflammatory phenotypes | 1.69 | 2.23 | <0.001 | *** |
| Eosinophilic vs No asthma | 1.06 | 2.93 | 0.003 | ** |
| Paucigranulocytic vs No asthma | 0.44 | 1.40 | 0.14 |  |
| Neutrophilic vs No asthma | 1.97 | 3.59 | <0.001 | *** |
| Mixed vs No asthma | 0.69 | 1.06 | 0.40 |  |
| Age | 1.19 | 6.30 | <0.001 | *** |
| Sex | 0.39 | 2.04 | 0.027 | * |
| Country | 6.60 | 8.72 | <0.001 | *** |

Significance codes: *** *P*<0.001, ** *P*<0.01, * *P*<0.05.

**Supplementary** **Table 5**: Permutational multivariate analysis of variance (PERMANOVA) output assessing the effect of neutrophil% and eosinophil% on microbiota composition, within country, adjusting for age and sex.#

|  | **R^2^ (%)** | **Pseudo-F** | ***P* value** | **Significance** |
| --- | --- | --- | --- | --- |
| **Brazil** |  |  |  |  |
| Neutrophil% | 2.96 | 1.81 | 0.057 |  |
| Eosinophil% | 3.08 | 1.88 | 0.051 |  |
| Age | 2.02 | 1.24 | 0.24 |  |
| Sex | 2.10 | 1.29 | 0.21 |  |
| **Ecuador** |  |  |  |  |
| Neutrophil % | 1.09 | 0.97 | 0.46 |  |
| Eosinophil% | 0.64 | 0.57 | 0.88 |  |
| Age | 2.76 | 2.46 | 0.005 | ** |
| Sex | 1.14 | 1.02 | 0.42 |  |
| **New Zealand** |  |  |  |  |
| Neutrophil% | 2.51 | 3.39 | <0.001 | *** |
| Eosinophil% | 0.10 | 1.36 | 0.17 |  |
| Age | 4.04 | 5.46 | <0.001 | *** |
| Sex | 0.68 | 0.92 | 0.50 |  |
| **Uganda** |  |  |  |  |
| Neutrophil% | 2.96 | 1.78 | 0.066 |  |
| Eosinophil% | 1.20 | 0.72 | 0.67 |  |
| Age | 0.89 | 0.64 | 0.79 |  |
| Sex | 1.50 | 0.92 | 0.48 |  |

Significance codes: *** *P*<0.001, ** *P*<0.01, * *P*<0.05.

#Analysis not performed for UK

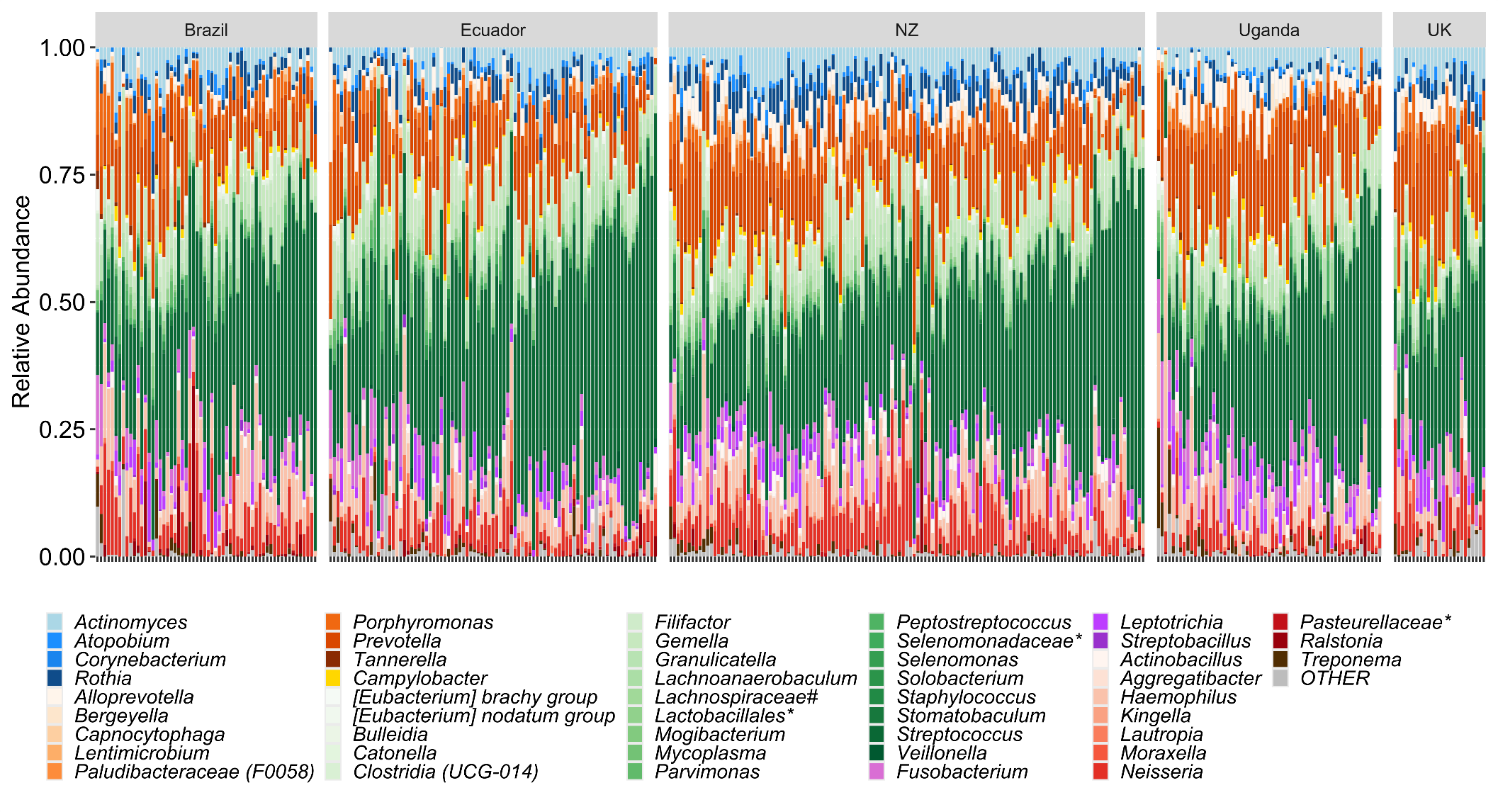

**Supplementary** **Figure 1: Taxa bar plot of WASP-biome.** Relative abundance distribution of taxa present in ≥10% of individuals within a country. Taxa coloured by phylogeny where blue = Actinobacteriota (n=4), orange = Bacteroidota (n=8), gold = Campilobacterota (n=1), green = Bacillota (n=22), purple = Fusobacteriota (n=3), red = Pseudomonadota (n=9), brown = Spirochaetes (n=1). * unassigned; # uncultured.

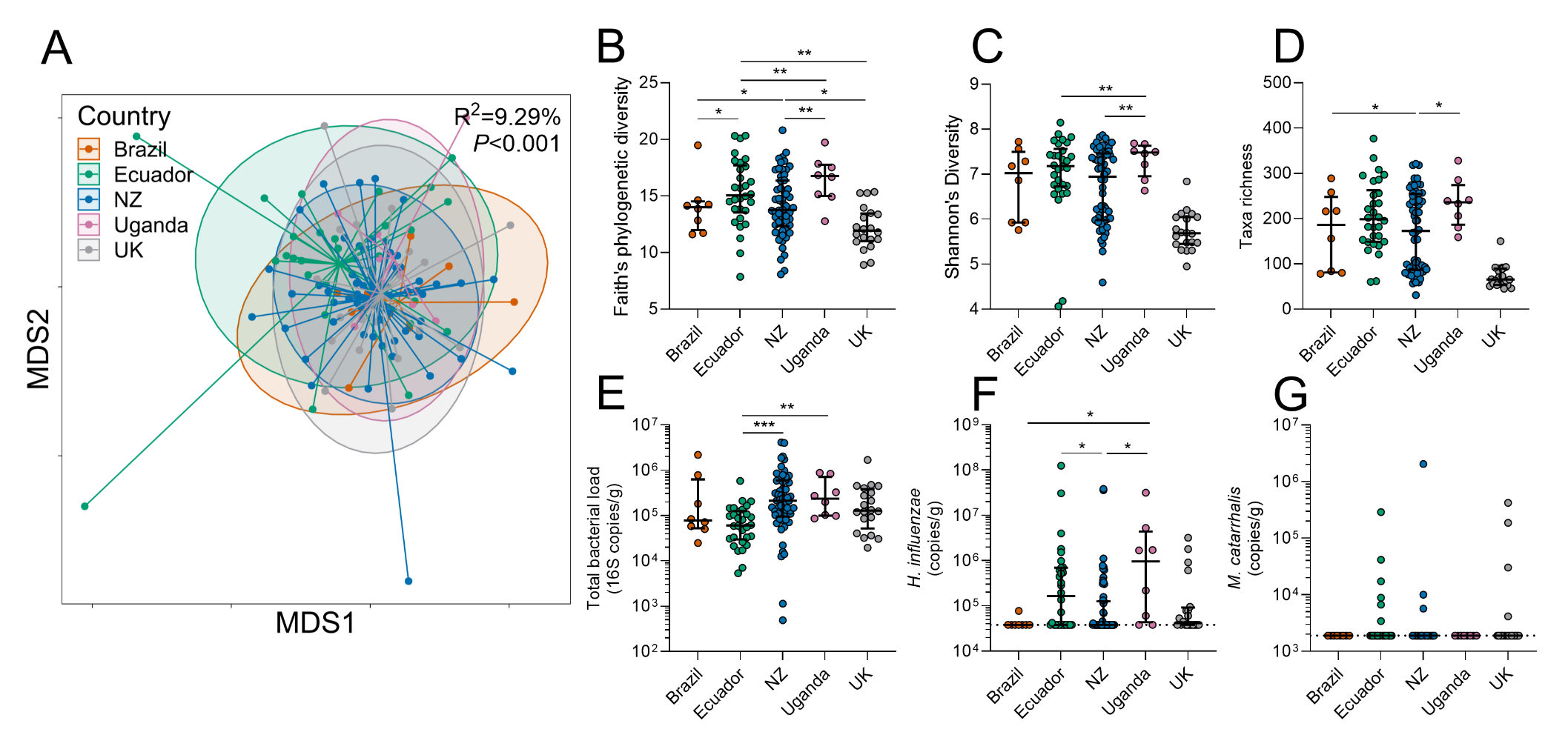

**Supplementary** **Figure 2: Sputum microbiota differs by country in non-asthmatics.** A) Nonmetric multidimensional scaling (NMDS) plot of Bray-Curtis dissimilarity. B) Shannon’s diversity. C) Faith’s phylogenetic diversity. D) Taxa richness. E) Total bacterial load derived from qPCR. F) *Haemophilus influenzae* abundance derived from qPCR. G) *Moraxella catarrhalis* abundance derived from qPCR. Statistics: A) Permutational multivariate analysis of variance including variables: country, age and sex; B-G) Ordinal logistic regression including variables: country, age and sex; *** *P*<0.001, ** *P*<0.01, * *P*<0.05.

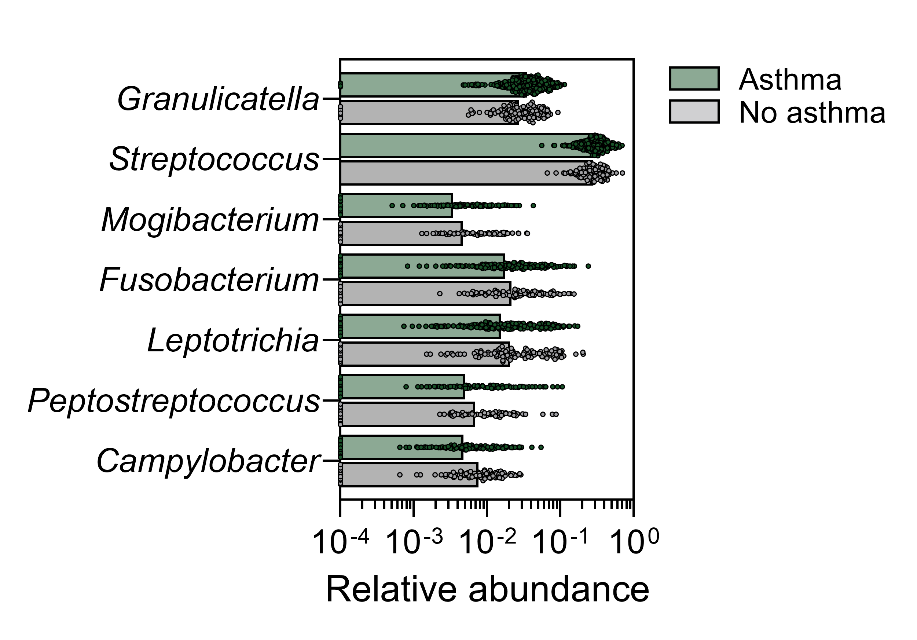

**Supplementary Figure 3: Taxa that differed significantly by asthma status.** Distribution of the relative abundance of taxa either significantly higher or lower in asthmatics. Analysis performed by ordinal logistic regression including variables: asthma, country, age and sex.

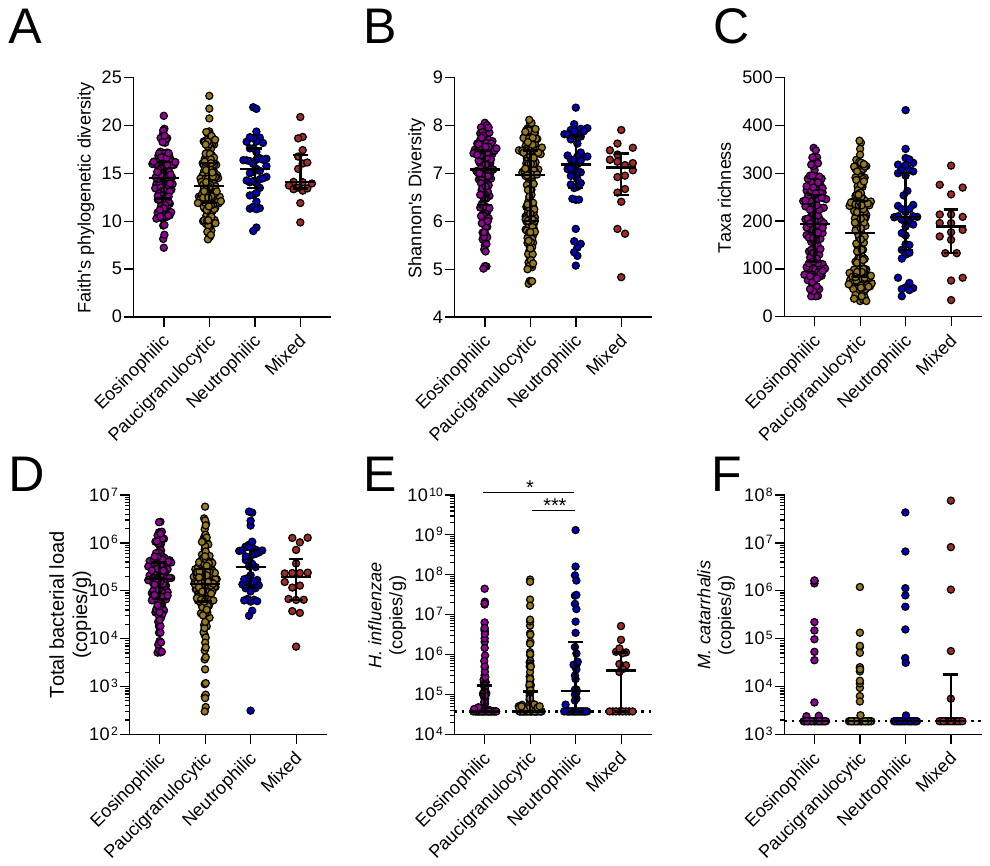

**Supplementary** **Figure 4:** **Sputum microbiota by inflammatory phenotype in asthmatics**. A) Faith’s phylogenetic diversity. B) Shannon’s diversity. C) Taxa richness. D) Total bacterial load derived from qPCR. E) *Haemophilus influenzae* abundance derived from qPCR. F) *Moraxella catarrhalis* abundance derived from qPCR. Statistics: Ordinal logistic regression including variables: inflammatory phenotype, country, age and sex; * *P*<0.05, *** *P*<0.001.

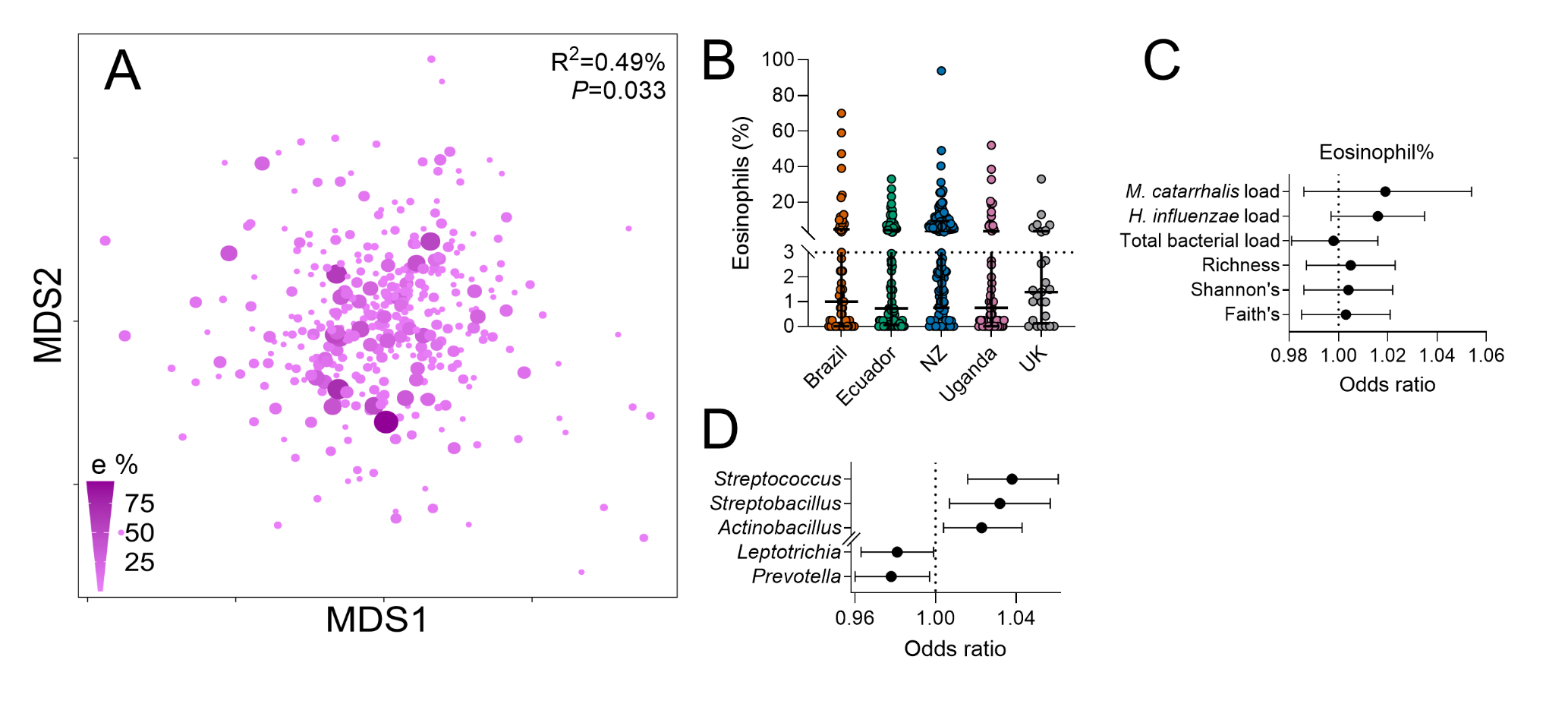

**Supplementary** **Figure 5: Sputum microbiota and eosinophilic phenotype in asthma.** A) Nonmetric multidimensional scaling (NMDS) plot of Bray-Curtis dissimilarity showing dispersion by eosinophil%. B) Distribution of sputum eosinophil% by country. C) Forest plot showing eosinophil% associated with α-diversity (Faith’s, Shannon’s, richness) and qPCR derived bacterial load (total, and *H. influenzae* and *M. catarrhalis* specific). D) Forest plot showing taxa that differed significantly by eosinophil%. Statistics: A) Permutational multivariate analysis of variance including variables: neutrophil%, eosinophil%, country, age and sex; C, D) Ordinal logistic regression including variables: neutrophil%, eosinophil%, country, age and sex.

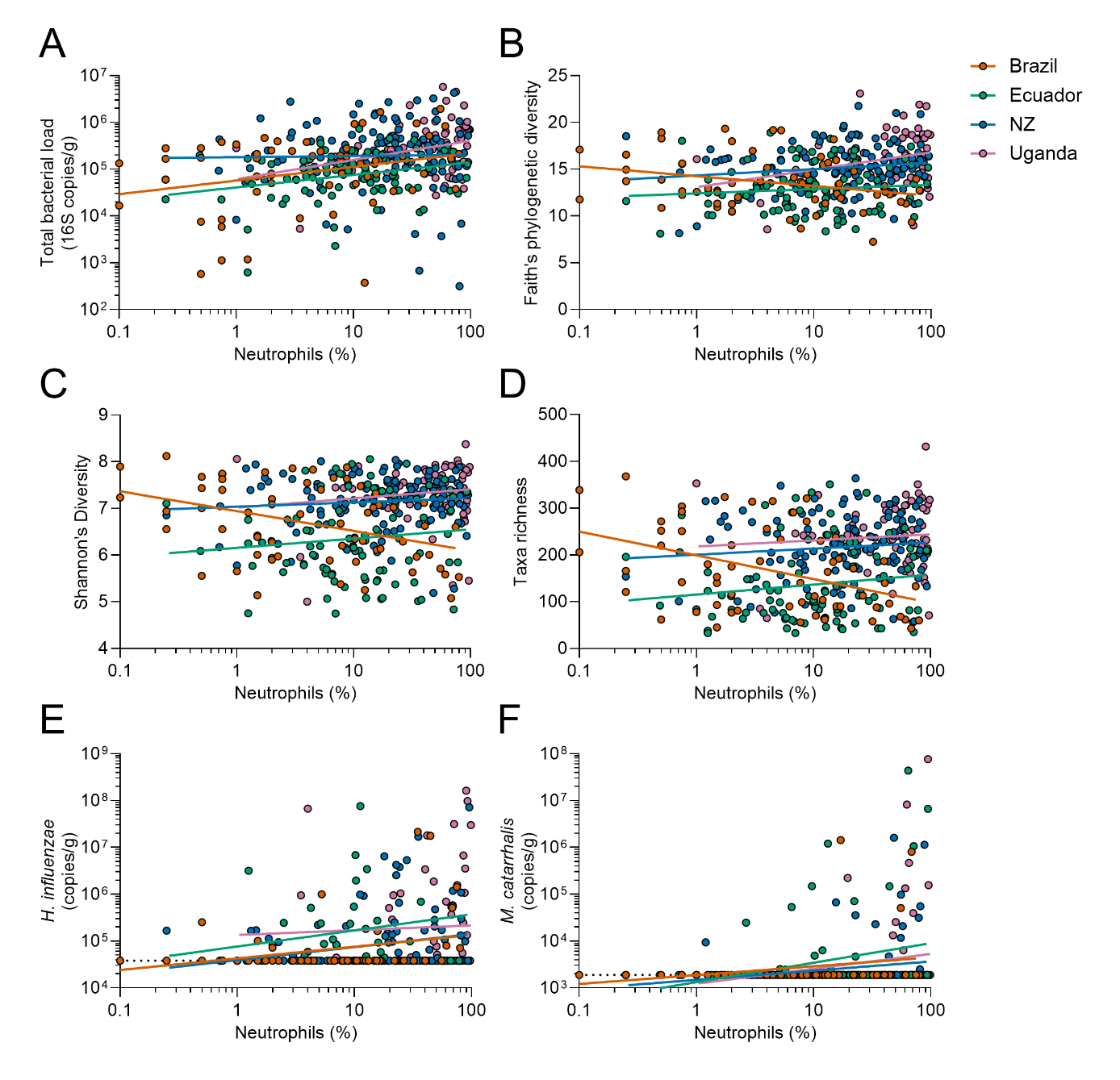

**Supplementary** **Figure 6: Within-country scatter plots between sputum microbiota characteristics and neutrophil%.** A) Total bacterial load derived from qPCR. B) Faith’s phylogenetic diversity. C) Shannon’s diversity. D) Taxa richness. E) *Haemophilus influenzae* abundance derived from qPCR. F) *Moraxella catarrhalis* abundance derived from qPCR. Linear regression lines included for visualisation. Analysis not performed for the UK due to low sample size.

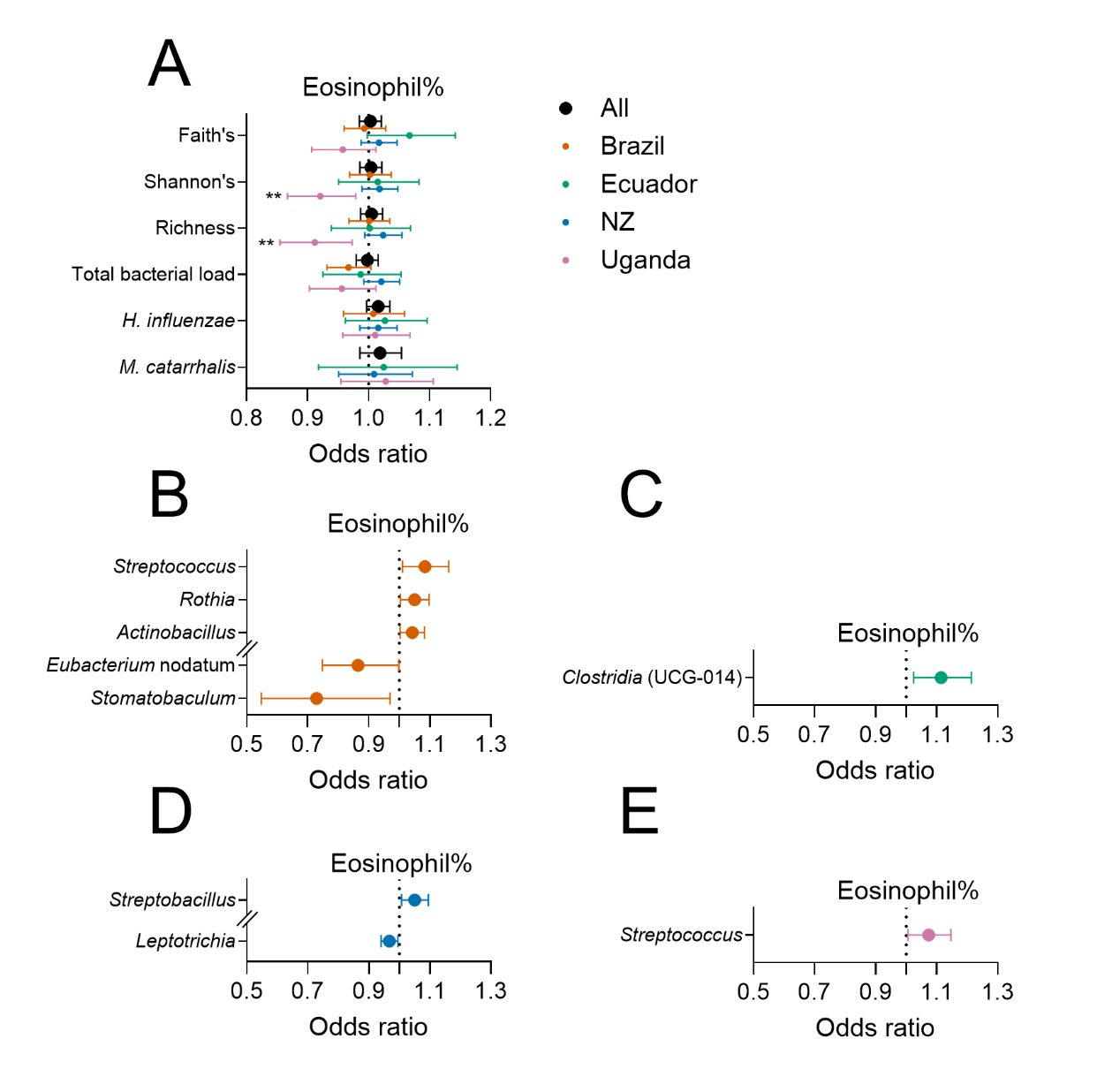

**Supplementary** **Figure 7: Within-country association between sputum microbiota and eosinophil%.** A) Forest plot showing eosinophil% associated with α-diversity (Faith’s, Shannon’s, richness) and qPCR derived bacterial load (total, and *H. influenzae* and *M. catarrhalis* specific) in whole cohort (black), and within Brazil (orange), Ecuador (green), New Zealand (blue), and Uganda (pink). B) Forest plot showing taxa that differed by eosinophil% in Brazil. C) Forest plot showing taxa that differed by eosinophil% in Ecuador. D) Forest plot showing taxa that differed by eosinophil% in New Zealand. E) Forest plot showing taxa that differed by eosinophil% in Uganda. Statistics: Ordinal logistic regression including variables: neutrophil%, eosinophil%, age and sex; *** *P*<0.001, ** *P*<0.01, * *P*<0.05. *M. catarrhalis* analysis was not performed for Brazil due to low detection frequency (3 out of 60 participants).
